# Interval timing and midfrontal delta oscillations are impaired in Parkinson’s disease patients with freezing of gait

**DOI:** 10.1101/2021.05.18.21257273

**Authors:** Taylor J. Bosch, Richa Barsainya, Andrew Ridder, KC Santosh, Arun Singh

## Abstract

Gait abnormalities and cognitive dysfunction are common in patients with Parkinson’s disease (PD) and get worst with disease progression. Recent evidence has suggested a strong relationship between gait abnormalities and cognitive dysfunction in PD patients and impaired cognitive control could be one of the causes for abnormal gait patterns. However, the pathophysiological mechanisms of cognitive dysfunction in PD patients with gait problems are unclear. Here, we collected scalp electroencephalography (EEG) signals during a 7-second interval timing task to investigate the cortical mechanisms of cognitive dysfunction in PD patients with (PDFOG+, n=34) and without (PDFOG–, n=37) freezing of gait, as well as control subjects (n=37). Results showed that the PDFOG+ group exhibited the lowest maximum response density at around 7 seconds compared to PDFOG– and control groups, and this response density peak correlated with gait abnormalities as measured by FOG scores. EEG data demonstrated that PDFOG+ had decreased midfrontal delta-band power at the onset of the target cue, which was also correlated with maximum response density and FOG scores. In addition, our classifier performed better at discriminating PDFOG+ from PDFOG– and controls with an area under the curve of 0.93 when midfrontal delta power was chosen as a feature. These findings suggest that abnormal midfrontal activity in PDFOG+ is related to cognitive dysfunction and describe the mechanistic relationship between cognitive and gait functions in PDFOG+. Overall, these results could advance the development of novel biosignatures and brain stimulation approaches for PDFOG+.

## Introduction

The association between cognitive function and gait disturbances or freezing of gait (FOG) in Parkinson’s disease (PD) has been studied in recent years [1-4]. Gait disturbances are a lower-limb motor symptoms of PD; however, cognitive dysfunction is a common non-motor complication of PD, with estimated point prevalence rates of more than 60% [5]. Cognitive dysfunction in PD is the most consequential non-motor feature of the PD, contributing to reduced quality of life and increased risk for disability and mortality [6]. A previous study has suggested that gait abnormalities can predict cognitive dysfunction in PD patients [7]. Several studies have shown deprived cognitive function in PD patients with FOG (PDFOG+) compared to those without FOG (PDFOG–), suggesting that the cognitive domains of executive function, attention, and visuospatial function are attenuated in PDFOG+ [8, 9]. There are currently no therapies that significantly improve both the cognitive and lower-limb motor/gait symptoms of PD because the neural mechanisms of both symptoms and the mechanistic relationship between cognitive and lower-limb motor impairments are unknown.

Cognitive dysfunctions in neurodegenerative and neuropsychiatric diseases can be explored via many executive tasks such as the interval timing task, which requires subjects to estimate temporal intervals of several seconds [10]. The interval timing task is a time perception task with temporal information processing and requires working memory for temporal rules and attention to the passage of time. Interval timing is thought to involve prefrontal cortical areas, which are associated with working memory and attention [11]. We have investigated the interval timing task in patients with PD and schizophrenia; and have observed abnormal behavioral responses compared to healthy controls [12, 13]. PD patients with gait impairments show further abnormal gait patterns when they execute a timed performance while walking, suggesting the presence of distorted time perception during dual tasks [14].

Midfrontal low frequency (delta;1-4 Hz/theta; 4-7 Hz) oscillations have been proposed as the mechanism of cognitive control in PD patients [15, 16]. Our prior scalp electroencephalography (EEG) studies have shown attenuated midfrontal delta/theta oscillations during executive cognitive functions in PD patients compared to control subjects [12, 16]. We have also observed reduced midfrontal theta activity at the occurrence of the ‘Go’ cue during lower-limb movements that require attention or active cognitive processing in PD patients with FOG (PDFOG+) compared to without FOG (PDFOG–) and controls [17]. Since gait requires active cognitive-motor dual task control [2, 17, 18], midfrontal low-frequency oscillations in cortical and associated networks may be critical in executing motor tasks with cognitive load and may be involved in the freezing phenomenon in PDFOG+ [16, 19]. However, the differences in the neural mechanisms of time perception or interval timing between PDFOG+ and PDFOG– are unclear. The current study sought to examine the differences in interval timing and the underlying neural mechanisms between PDFOG+ and PDFOG–.

## Materials and methods

### Participants and clinical assessments

A total of 109 participants including 74 patients with PD (34 PDFOG+ and 37 PDFOG–) and 37 age matched healthy controls performed the interval timing task. All procedures were approved by the Institutional Review Board. Most of the datasets have been analyzed previously to investigate the interval timing deficits in PD patients and control subjects [12], however, in previous report, PDFOG+ and PDFOG– groups were not investigated.

Clinical demographics are demonstrated in Table 1. Disease severity and FOG in PD patients were assessed via the motor portion of the Unified Parkinson’s Disease Rating Scale (mUPDRS) [20] and the FOG questionnaire [21], respectively. PDFOG+ were selected on the basis of the following criteria: a) patients confirmed they had difficulty in starting, stopping, and turning during a movement; b) their FOGQ score was greater than zero, suggestive of at least one FOG episode in the past month; c) moreover, PDFOG+ were re-identified by a movement disorders specialist; d) an objective confirmation of PDFOG+, an unassisted walk with rapid turning, was performed prior to the study. All participants were assessed in the ‘ON’ anti-parkinsonian medication state.

**Table 1.** Demographic and clinical assessments.

| Measure | Control<br>(n=37) | PD<br>(n=71) | PDFOG–<br>(n=37) | PDFOG+<br>(n=34) | Control vs.<br>PD | PDFOG– vs.<br>PDFOG+ |
| --- | --- | --- | --- | --- | --- | --- |
|  |  |  |  |  | Independent<br>t-test | Independent<br>t-test |
| Gender (M/F) | 21/16 | 50/21 | 25/12 | 25/9 | 1.38 | 0.54 |
| Age (years) | 72 ± 7 | 69 ± 8 | 68.4 ± 7.9 | 70.0 ± 8.2 | 2.04 (0.04)* | -0.31 (0.76) |
| DD (years) | - | 5 ± 4 | 4.4 ± 3.3 | 5.8 ± 4.6 | - | -1.52 (0.13) |
| LEDD (mg) | - | 848 ± 462 | 732 ± 408 | 973 ± 491 | - | -2.26 (0.03)* |
| FOG | - | 6.3 ± 5.7 | 1.7 ± 1.3 | 11.3 ± 4.1 | - | -13.22 (<0.001)*** |
| mUPDRS | - | 12.8 ± 7 | 9.0 ± 5.4 | 17.0 ± 6.2 | - | -5.79 (<0.001)*** |

DD: Disease duration; LEDD: Levodopa equivalent daily dose; FOG: Freezing of Gait; mUPDRS: motor
Unified Parkinson's Disease Rating Scale.
Presented as t-value (p-value). \* p<0.05; \*\*\*p<0.001

The interval timing task has been described in our previous reports [12, 13]. In summary, to measure cognitive function, all participants performed either 7-second or 3-second interval timing trials [12, 13]. Patients were instructed with the text “short interval” and “long interval” for 3-second and 7-second interval timing trials, respectively. After instructions, a target cue appeared on the computer screen. Participants pressed the spacebar when they thought that 3 seconds for short or 7 seconds for longer interval timing trials had been elapsed. All participants performed 6 training trials and the experiments consisted of 40 randomized trials of each interval trial type (20 trials/block, total 4 blocks). In our main study, we did not observe any significant changes in behavioral responses and midfrontal activity during the 3-second interval timing task between PD patients and control subjects [12]. Therefore, in the current study, we only selected the 7-second interval timing trials for the analyses (Fig. 1A). We analyzed the mean response time of the keypress time for timing accuracy of participants. We used the Matlab “ksdensity” function to compute the probability density estimate of response time and plot the data, where the estimate was based on a normal kernel function. Additionally, we selected the 6.5 to 7.5 seconds window to compute maximum response density values.

**Fig. 1.**
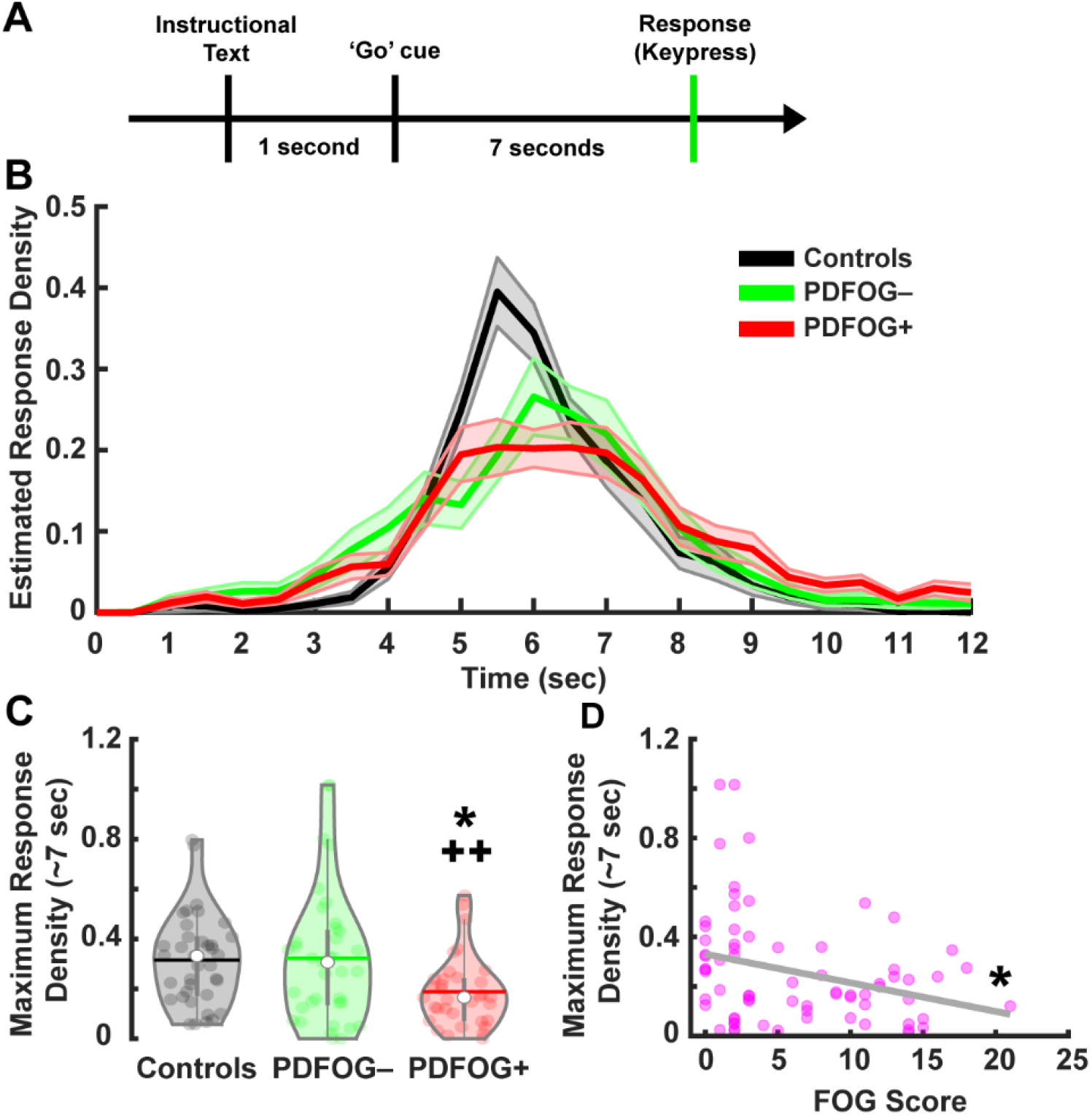
PDFOG+ exhibit a decreased ability to consistently estimate timing (as measured by the maximum response density at ∼7 seconds). (A) Overview of the 7-second interval timing trial. Participants received a ‘Go’ cue and responded with a keypress when they estimated the target time (7 seconds) had elapsed. (B) Estimated response density distribution for response time plotted from 0-12 seconds from the ‘Go’ cue. (C) PDFOG+ exhibited reduced maximum response density between 6.5 and 7.5 seconds compared to PDFOG– and control groups. (D) Maximum response density between 6.5 and 7.5 seconds was also correlated with gait abnormalities assessed by the freezing of gait questionnaire (FOGQ). *p < 0.05 vs. PDFOG–. ++p < 0.01 vs. controls. *p < 0.05 significant correlation.

### Scalp EEG recording and analysis

We used a 64-channel customized EEG cap and the Brain Vision amplifier (Brain Products GmbH) to collect signals with a 0.1 Hz high-pass filter and a 500 Hz sampling rate. Electrodes Pz and FPz were used as a reference and ground, respectively, similar to our previous report [12]. We removed unreliable Fp1, Fp2, FT9, FT10, TP9, and TP10 channels before preprocessing the signals, resulting in 59 channels. Signals were segmented around the onset of the instructional text cue (−2 s to 20 s for 7-second interval timing trials) and then the target ‘Go’ cue-triggered and corresponding response-triggered segments were isolated for further analysis. Signal from the reference electrode was recovered using the average reference method. Bad channels and bad epochs were identified using the FASTER and pop_rejchan algorithms with default parameters [22]. Eye movement and other motor-related artifacts were removed using independent component analysis. Since our *a priori* hypothesis was focused on midfrontal low-frequency bands, we focused our analyses on the midfrontal Cz electrode in the current study to be consistent with our prior reports [12, 16, 23-27].

Similar to our previous reports [12, 13, 16, 17], we performed time-frequency analyses on each trial using the Morlet wavelets method (defined as a Gaussian-windowed complex sine wave: e^i2πtf^e^-t^2/(2xσ^2)^, where *t*=time and *f* =frequency) [28] and averaged to estimates of instantaneous power. We focused the analysis on the -500 – +1000 ms time window for both cue-triggered and response-triggered trials. Power was normalized by converting to a decibel (dB) scale (10*log10(power_t_/power_baseline_)), to compare the effects across frequencies. Similar to our previous reports, the baseline for each frequency was computed by averaging power from □300 – -200 ms prior to the occurrence of the ‘Go’ cue [16, 26]. The time-frequency regions of interest (tf-ROIs) were restricted to pre-selected frequency bands for cue-triggered trials: delta-band (1-4 Hz) and theta-band (4-7 Hz). For response-triggered trials, we included the beta-band (13-21 Hz) as well [16, 23-27, 29-31]. For the cue- and response-triggered trials, we selected 0 – 500 ms and -250 – 250 ms for the tf-ROIs, respectively. In addition to this well-motivated tf-ROI, we performed a cluster-based permutation correction to time-frequency data with a cluster size of 500 pixels and an independent t-test to detect any other reliable changes between groups [23, 26].

### Statistical analyses and classification

All clinical assessments were compared between PD (including both PDFOG+ and PDFOG–) and controls, as well as between PDFOG+ and PDFOG–, using independent *t-tests*. We used one-way analysis of variance (ANOVA) followed by pairwise comparisons with Bonferroni corrections to compare behavioral and EEG data across groups (controls, PDFOG–, and PDFOG+). Eta squared was used to measure the effect size for ANOVAs; and Cohen’s d was determined to compute effect size for pairwise comparisons. Pearson’s correlation tests were implemented for all correlation analyses. Further, mediation analysis was performed to determine if disease severity (mUPDRS score) mediates the relationship between gait abnormalities (FOG score) and cue-triggered delta power. The Sobel test was performed to calculate the significance of a mediation effect. All values are shown in mean ± standard deviation.

To construct an inclusive model, we applied linear mixed-effects modeling tests in Matlab using the ‘fitlme’ function. Maximum response density at ∼7 sec and cue-triggered tf-ROIs (delta and theta power values) were response variables and FOG score was the predictor variable. The grouping variable included controls, PDFOG–, and PDFOG+.

In addition, we performed classification analyses (three times) to classify PDFOG+ from PDFOG– and controls on the basis of cue-triggered delta-band, theta-band, and including both bands in Matlab (MathWorks) using the Machine-learning toolbox. For multiclass classification, we used the ‘fitctree’ function to get a fitted binary classification decision tree. Then ‘resubPredict’ function returned the labels that the model predicted for the training data. We computed the receiver operating characteristic (ROC) curve using the ‘perfcurve’ function for the predictions that an observation belonged to PDFOG+, given the true class labels participants; and computed the optimal operating point and area under the curve (AUC). ROC graphs represented the performance of the classification models and plotted true positive rate against false positive rate.

## Results

### Behavioral outcomes

The estimated probability density distribution for response times can be visualized in Figure 1B. To assess the probability of responding at the targeted time (7-second), we compared the maximum response density between 6.5 and 7.5 seconds across groups. The results from the one-way ANOVA for maximum response density showed a main effect of group (F_2,105_ = 4.87, p = 0.009, η^2^ = 0.085; Fig. 1C). Subsequent pairwise comparisons demonstrated a difference between PDFOG+ (0.19 ± 0.02 maximum response density) and controls (0.32 ± 0.03 maximum response density) (p = 0.002, d = -0.78) as well as PDFOG+ and PDFOG– (0.32 ± 0.04 maximum response density) (p = 0.01, d = -0.63). No difference was demonstrated between PDFOG– and controls (p = 0.88, d = 0.04). Pearson correlation analyses showed a significant association between maximum response density and FOG scores (r^2^ = -0.29, p = 0.013; Fig. 1D). Similar to our prior report [12], we found no effect of group when assessing mean response time (F_2,105_ = 1.8, p = 0.17, η^2^ = 0.033; Fig. S1A), though a significant correlation was observed between mean response time and FOG scores (r^2^ = -0.29, p = 0.01; Fig. S1B). Further, clinical correlational analyses revealed significant associations between disease severity (as measured by the mUPDRS) and FOG scores (r^2^ = 0.63, p < 0.001). Overall, these results suggest a striking difference in the ability of PDFOG+ to make consistent timing predictions on a trial-by-trial basis which are not reflected when examining overall performance across an entire testing session.

### Midfrontal low-frequency oscillatory activity

To examine the neurophysiological manifestations of PDFOG+, we examined low-frequency delta- and theta-band oscillations in response to the presentation of the ‘Go’ cue in the 7-second interval timing task (Fig. 2A and B). The results from the one-way ANOVA for cue-triggered delta-band power showed a main effect of group (F_2,105_ = 4.27, p = 0.016, η^2^ = 0.075; Fig. 2C). Subsequent pairwise comparisons demonstrated a difference between PDFOG+ (0.22 ± 0.15 dB) and controls (0.49 ± 0.19 dB) (p = 0.005, d = -0.69) as well as between PDFOG+ and PDFOG– (0.27 ± 0.18 dB) (p = 0.04, d = -0.5). No difference was demonstrated between PDFOG– and controls (p = 0.39, d = -0.2). Correlation analysis showed a significant association between FOG scores and cue-triggered delta-band power (r^2^ = -0.24, p = 0.047; Fig. 2D) as well as an association between maximum response density and cue-triggered delta-band power (r^2^ = 0.43, p < 0.001; Fig. 2E). Time-frequency plots for the complete time window can be visualized for each group in Figure S2. Since we observed a clinical correlation between FOG scores and disease severity (mUPDRS), we also performed a mediation analysis to assess whether disease severity mediated the relationship between gait abnormalities (FOG score) and cue-triggered delta-band power. Mediation analysis revealed no indirect influence of disease severity on delta-band power (r^2^ = 0.08, Sobel test: p = 0.25), indicating that disease severity did not drive an illusory correlation between FOG scores and cue-triggered delta-band power.

**Fig. 2.**
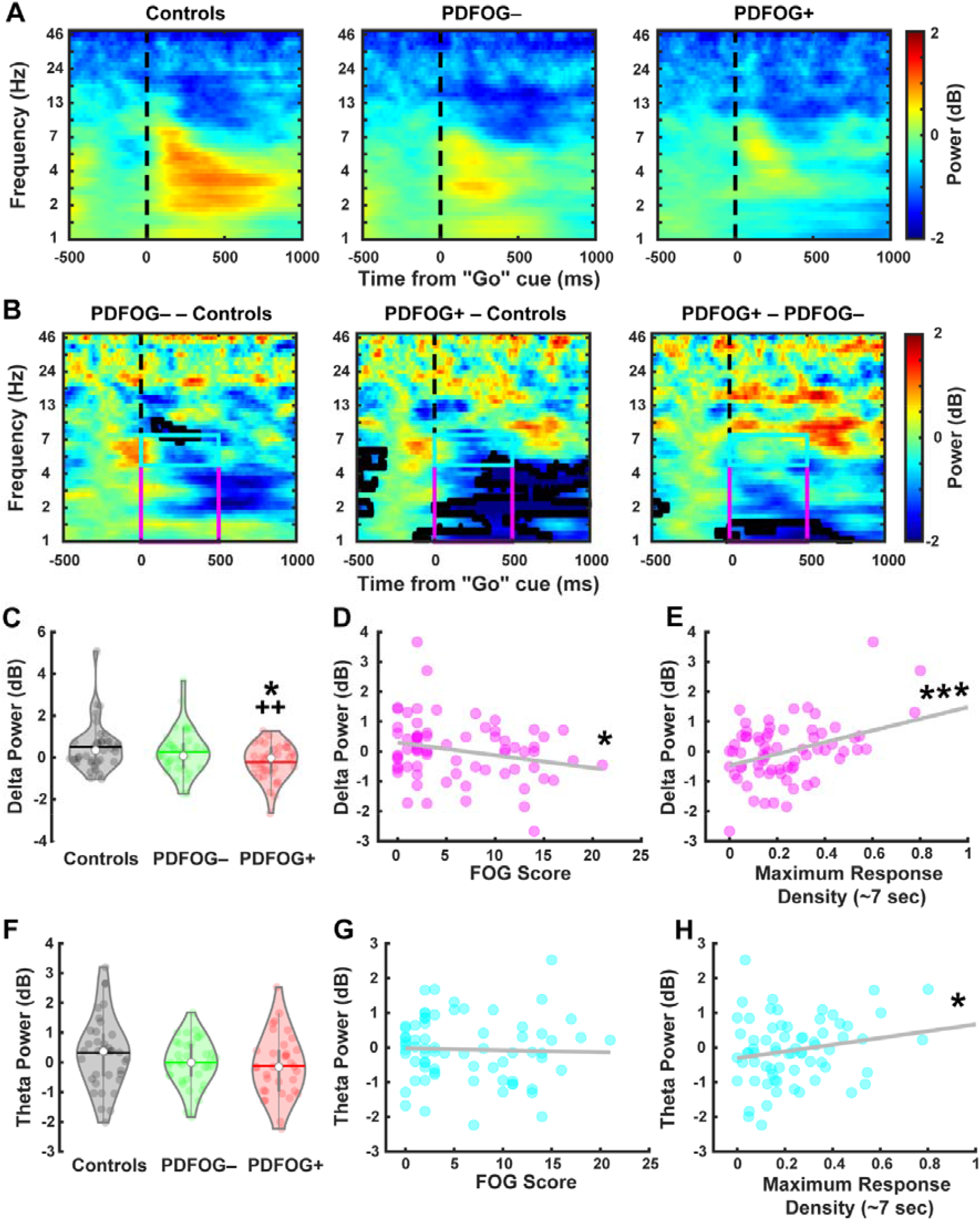
PDFOG+ show decreased cue-triggered midfrontal delta-band power during the interval timing task. (A) Time-frequency distribution in controls, PDFOG–, and PDFOG+ during the interval timing task. (B) Time-frequency analyses comparing PDFOG+, PDFOG-, and controls. (C) Delta power (1-4 Hz, tf-ROI: red box) was significantly decreased in the PDFOG+ group compared to PDFOG– and control groups. (D) Gait abnormalities assessed by the freezing of gait (FOG) score in PD patients were correlated with delta power during the interval timing task. (E) Maximum response density at ∼7 seconds was also associated with delta power during the interval timing task. (F) However, theta power (4-7 Hz, tf-ROI: cyan box) was not different in PDFOG+ compared to other groups. (G-H) Gait abnormalities and maximum response density in PD patients were not correlated with theta power. *p < 0.05 vs. PDFOG–. ++p < 0.01 vs. controls. *p < 0.05 significant correlation. ***p < 0.001 significant correlation. B: Areas outlined by solid black lines indicate p < 0.05 via a t-test.

The results from the one-way ANOVA for cue-triggered theta-band power showed no main effect of group (F_2,105_ = 1.73, p = 0.18, η^2^ = 0.03; Fig. 2F). Pearson correlation analysis showed no association between FOG scores and cue-triggered theta-band power (r^2^ = -0.04, p = 0.77; Fig. 2G), though an association was seen between maximum response density and cue-triggered theta-band power (r^2^ = 0.24, p = 0.048; Fig. 2H). Overall, these data suggest that neurophysiological deficits in PDFOG+ manifest in the low-frequency delta-band in response to the ‘Go’ cue. Furthermore, the associations between FOG scores, delta-band power, and maximum response density support a strong link between behavioral deficits seen in PDFOG+ and low frequency midfrontal neural oscillations.

In addition to cue-triggered EEG activity, we also sought to explore EEG responses at the onset of movement or keypress. Differences in response-triggered EEG activity can be visualized in Figure S3A. The results from the one-way ANOVAs for response-triggered delta-, theta-, and beta-band power revealed no main effects of group for delta-band power (F_2,105_ = 1.55, p = 0.22, η^2^ = 0.029; Fig. S3B), theta-band power (F_2,105_ = 2.31, p = 0.10, η^2^ = 0.04; Fig. S3C), or beta-band power (F_2,105_ = 2.11, p = 0.13, η^2^ = 0.04; Fig. S3D). These results suggest that the delta-band differences observed between PDFOG+ and PDFOG– in the cue-triggered EEG activity are specific to the presentation of the ‘Go’ cue and do not differ once movement execution is initiated. However, response-triggered decreased power in beta-band was significantly associated with the gait impairment in PD patients (r^2^ = -0.31, p = 0.01; Fig. S3D).

### Mixed modeling results

The results from linear mixed effect modeling using FOG scores as a predictor variable and controls, PDFOG–, and PDFOG+ as the grouping variable showed a significant contribution of FOG to the model predicting maximum response density at ∼7 seconds (p = 0.003) and cue-triggered delta power (p = 0.006), but no contribution to cue-triggered theta power (p = 0.23) (Table 2). These findings lend further support to the association between FOG scores and behavioral responses as well as the relationship between low-frequency delta oscillations and FOG.

**Table 2.** Linear Mixed Effects Modeling.

| **Fixed Effects** | **Estimate** | **SE** | **t-value** | **p-value** |
| --- | --- | --- | --- | --- |
| *Maximum Response Density at ~7-second* |  |  |  |  |
| Intercept | 0.322 | 0.024 | 13.292 | <0.001*** |
| FOG | -0.011 | 0.004 | -3.047 | 0.003** |
| *Cue-Triggered Delta Power* |  |  |  |  |
| Intercept | 0.401 | 0.125 | 3.204 | 0.002** |
| FOG | -0.052 | 0.018 | -2.808 | 0.006** |
| *Cue-Triggered Theta Power* |  |  |  |  |
| Intercept | 0.163 | 0.123 | 1.33 | 0.19 |
| FOG | -0.022 | 0.018 | -1.21 | 0.23 |

FOG: Freezing of Gait. \*\* p<0.01; \*\*\*p<0.001

### Classification

At the end, we used machine learning classification to determine whether cue-triggered delta-band power can reliably classify PDFOG+ from PDFOG– and control groups. After computing and plotting the ROC for true-positive vs. false-positive, the area under the curve (AUC) for cue-triggered delta-band power was 0.93 (Fig. 3A). In contrast, both cue-triggered theta band power alone (AUC = 0.81; Fig. 3B) and cue-triggered delta- and theta-band power combined (AUC = 0.90; Fig. 3C) performed poor classification. This indicates that cue-triggered delta-band power is sufficient to classify or predict PDFOG+ with high accuracy.

**Fig. 3.**
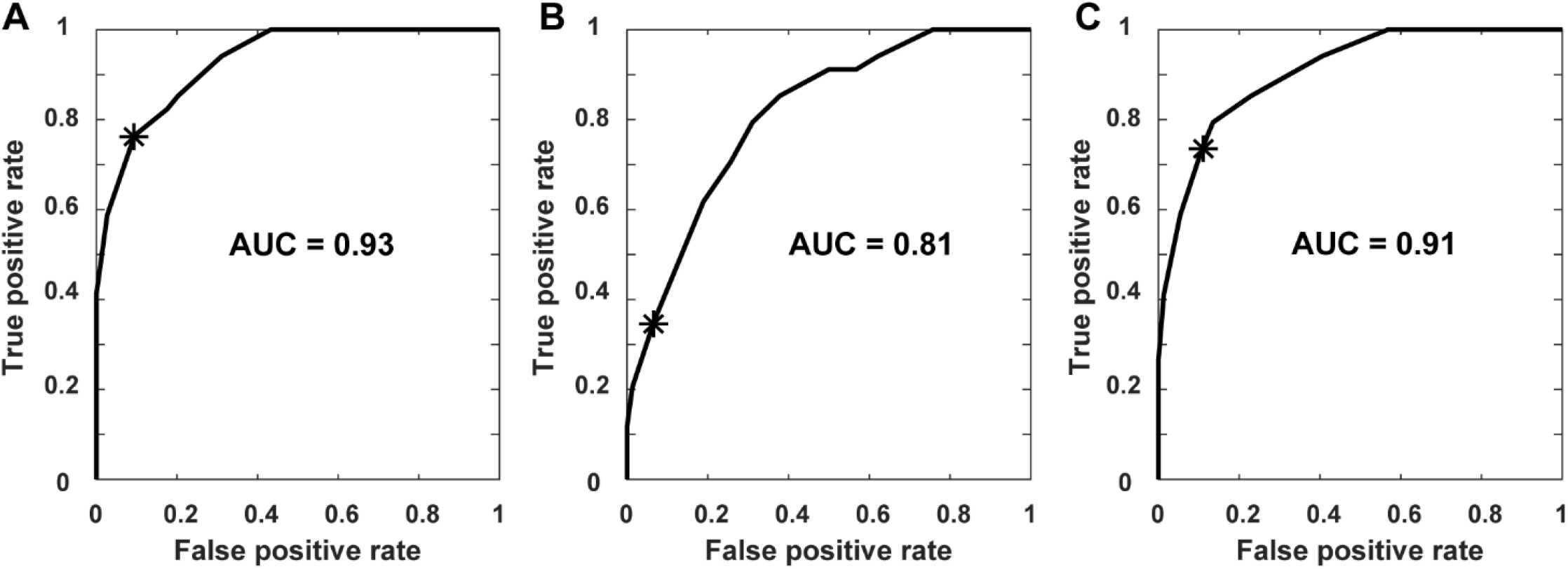
Cue-triggered midfrontal delta-band power is sufficient to classify PDFOG+. Machine learning classification was used to compute receiver operating characteristic (ROC) curves to classify PDFOG+ using (A) cue-triggered delta-band power, (B) cue-triggered theta-band power, and (C) cue-triggered delta- and theta-band power combined. Area under the curve (AUC) values represent classification accuracy with 1.0 being perfect classification. Asterisks represent the optimal operating point.

## Discussion

Current results demonstrate that PDFOG+ exhibit impaired time perception characterized by alterations in interval timing and midfrontal low-frequency delta-band oscillations. We observed that PDFOG+ performed the interval timing task with the lowest maximum response density at the 7-second interval timing task which correlated with gait impairment. Our results support previous studies demonstrating a significant relationship between gait impairment and deficits in cognitive function in PDFOG+ [1, 32, 33]. Our classification data also illuminated that interval timing task-related midfrontal low-frequency delta-band power can be used as a significant feature to classify PDFOG+. Certainly, current results suggest that PD patients with gait abnormalities exhibit higher timing deficits and dysfunctional midfrontal delta oscillations which are also associated with cognitive impairment. In addition, we observed a significant relationship between midfrontal delta power and gait abnormalities, and the mediation analysis showed no role of disease severity on that relationship, suggesting that cognitive deficits relate to severity of gait impairments. Altogether, these findings propose a cortical mechanism of cognitive dysfunction in PDFOG+.

Different models have been proposed regarding the location of pathology in the cortical and subcortical regions of PD patients with gait abnormalities and cognitive impairment [34, 35]. According to these models, any pathological or cellular mechanisms that impair the processing of cognitive control and interfere with temporal signaling or time perception may lead to an increase in gait abnormalities in PD. All these pathological alterations cause abnormal neurophysiological modulations, which can be identified by observing oscillatory activities in the cortical and subcortical regions. Abnormal gait patterns are associated with significant changes in subthalamic nucleus (STN) activity; and an association between abnormal STN activity and cognitive impairment has been seen in advanced PD patients, suggesting a critical role of the STN in gait and cognitive performances which can be targeted for deep brain stimulation therapy to improve both motor and cognitive functions in PDFOG+ [19, 27]. Our results are noteworthy in advancing the neurophysiological understanding of cognitive contributions to gait abnormalities in PDFOG+ because we demonstrate strong correlations between midfrontal delta power and both cognitive deficits (as measured by maximum response density at ∼7 sec) and gait dysfunction (as measured by FOG score). These findings suggest that cognitive dysfunction in PDFOG+ can result in part from their inability to recruit cognitive control processes, and that can be further impaired when PDFOG+ perform lower-limb movement/gait with cognitive load [36]. So, our data propose a cortical model in which significant alterations in delta-band activity contribute to cognitive dysfunction in PDFOG+.

These findings also provide evidence of time perception deficits or absent temporal information processing in PDFOG+. Previous studies have shown that PD patients show impaired working memory processing during interval timing as a function of dopamine signaling and that there can be further changes with disease progression such as in the PDFOG+ group [37, 38]. A previous imaging study showed that PD patients have a tendency to overestimate or underestimate intervals and suggested that dopamine may allow compensatory activation of the precuneus and consequently improve time estimation [39]. In addition to this, another study showed that a levodopa-treated PD group exhibited higher, faster pulse perception compared to untreated PD and control groups [40]. These findings are in agreement with the previous report that levodopa or dopamine intake does not improve interval timing deficits in PD patients [12]. Altogether, our results show that the occurrence of gait abnormalities with disease progression and increased dopamine deficits caused a delay in the internal clock and an under- or over-estimation of time in PDFOG+.

In general, the pathophysiology of severe timing deficits in PDFOG+ has been described as a dysfunction in the basal ganglia, which is a multisensory integration station that can explain time perception or temporal forecasting [41-43]; by contrast, our current and prior work strongly implicate cortical cognitive control mechanisms in PD [12, 16]. Studies have shown that cortical neurons participate in temporal processing and can be coherent at low-frequency delta-band oscillations [24, 44]. Such low-frequency delta oscillations can be seen in both cortical and subcortical neurons [27, 45]. Our data suggest that in PD patients with gait and cognitive dysfunctions, delta power is significantly decreased and results in inefficient engagement of temporal processing by fronto-striatal circuits [34, 46].

Stimulation at low frequencies can improve cognitive processing in PD patients [27, 47] and animal models of PD [24, 44], as well as in patients with other cognitive disorders such as schizophrenia [13], supporting the current results. Low-frequency STN stimulation has revealed an improvement in interval timing task performance in PD patients, most-likely via a prefrontal-subthalamic pathway [27]. Even anatomical studies clearly indicate that the basal ganglia nuclei participate in multiple circuits or ‘loops’ with cognitive areas of the cortical region [3, 41, 48]. Furthermore, STN deep brain stimulation at a high-frequency (130 Hz) improves upper-limb motor symptoms and cortical-STN beta power but may not affect cognitive processing in PD [49]. However, low-frequency 60 Hz STN [50] and 10–25 Hz pedunculopontine nucleus stimulation [51] can improve gait abnormalities in PDFOG+. Together, it appears that low-frequency stimulation at the basal ganglia nuclei might have potential to improve not only midfrontal abnormal low-frequency oscillations but also cognitive and gait performances in PDFOG+. Moreover, while our classifier method classifies PDFOG+ on the basis of midfrontal low-frequency delta power with higher accuracy, this midfrontal delta abnormality may not be highly predictive of PDFOG+. However, our results suggest that alterations in interval timing performance and midfrontal delta oscillations are predictive of cognition dysfunction in PDFOG+.

The current report has some limiting factors, which warrant consideration. First, even though EEG data contain low spatial resolution, data collected from combined EEG and fMRI or implanted deep brain stimulation electrodes might provide more detailed extrapolations about the alterations in cortical and subcortical regions during interval timing processing or cognitive functions in PDFOG+ compared to PDFOG–. Second, although not certainly a limiting factor for the current study, both PDFOG+ and PDFOG– groups performed the interval timing task while ‘on’ medication and therefore our results might be difficult to compare with previous studies. However, prior reports have shown no effects of levodopa on midfrontal low-frequency oscillations or cognitive behavioral performances such as interval timing or Simon reaction time tasks in PD patients [12, 16]. Third, the interval timing task contains both cognitive and keypress-related motor components. Our statistical analyses showed no changes in response-triggered low- and high-frequency oscillations between PDFOG+, PDFOG–, and controls. However, we observed that decreases in response-triggered beta-band power were related to gait impairment in PD patients [52].

In summary, the current study suggests that PDFOG+ exhibit higher cognitive deficits when performing temporal tasks such as interval timing which can manifest in altered midfrontal low-frequency delta-band activity. Both abnormal timing performance as measured by maximum response density at ∼7 seconds and midfrontal delta power show a significant relationship with gait impairment in PD. Overall, this report provides mechanistic insight into cognitive dysfunction and the relationship between cognitive impairment and gait abnormalities in PD.

## Data Availability

Data and analyses codes will be available on reasonable request.

## Data accession

The data and analysis codes will be available from the corresponding author upon reasonable request.

## Author contributions

TJ Bosch, R Barsainya, A Ridder, KC Santosh and A Singh conceived the project. TJ Bosch, R Barsainya, and A Singh performed the analyses. TJ Bosch, R Barsainya, A Ridder, KC Santosh and A Singh wrote and reviewed the manuscript.

## Acknowledgment

We thank Prof. Nandakumar Narayanan for providing required details about the dataset. This research was supported in part by the CBBRe research enhancement pilot grant program at the University of South Dakota.

## Supplementary data

Supplementary data related to this article can be found at

## Supplementary Information

**Fig. S1.**
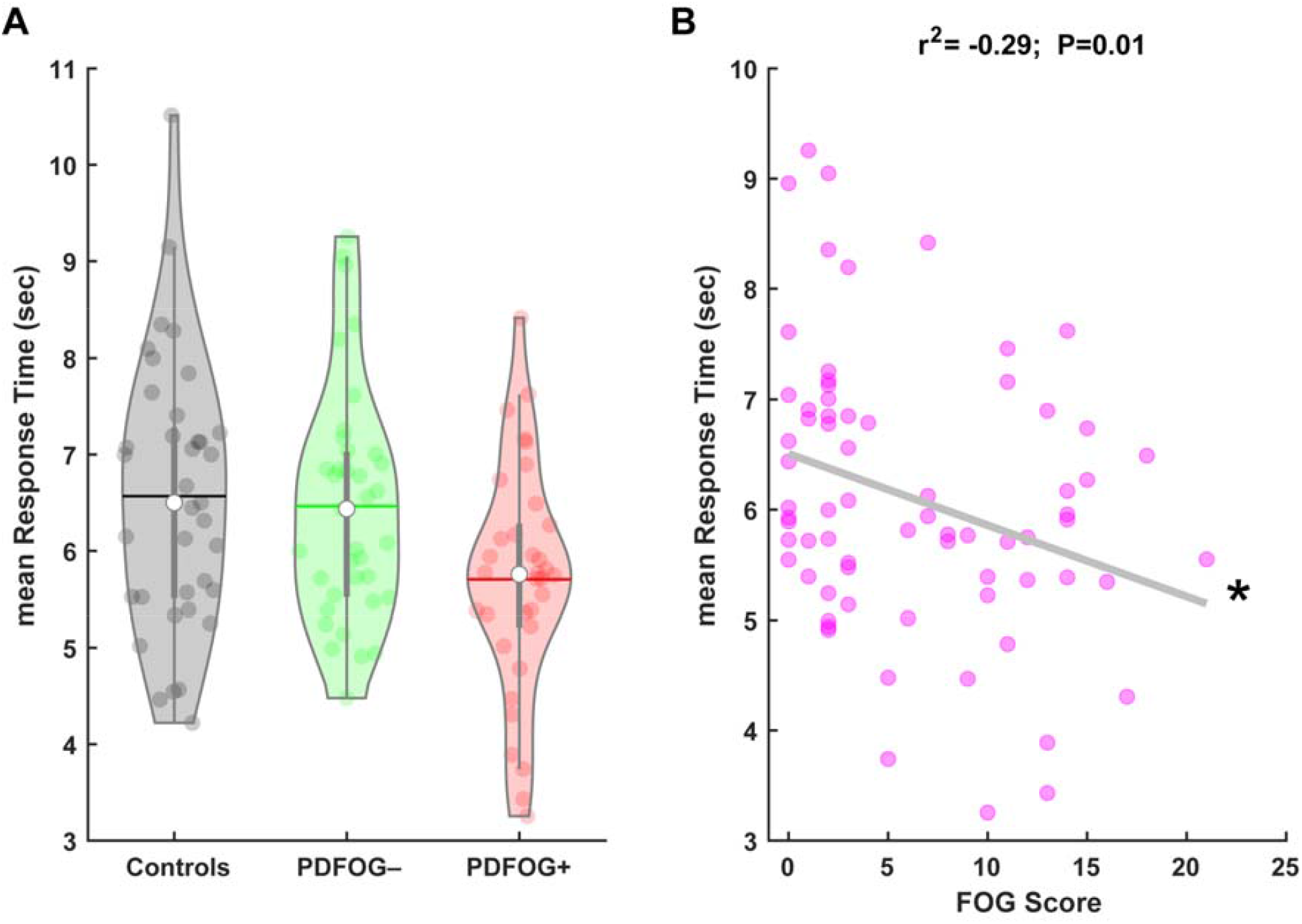
Behavioral results for mean reaction time during the 7-second interval timing task. (A) No difference in mean reaction time was observed across groups. (B) However, mean reaction time was associated with gait abnormalities assessed by the freezing of gait (FOG) score. *p < 0.05 significant correlation.

**Fig. S2.**
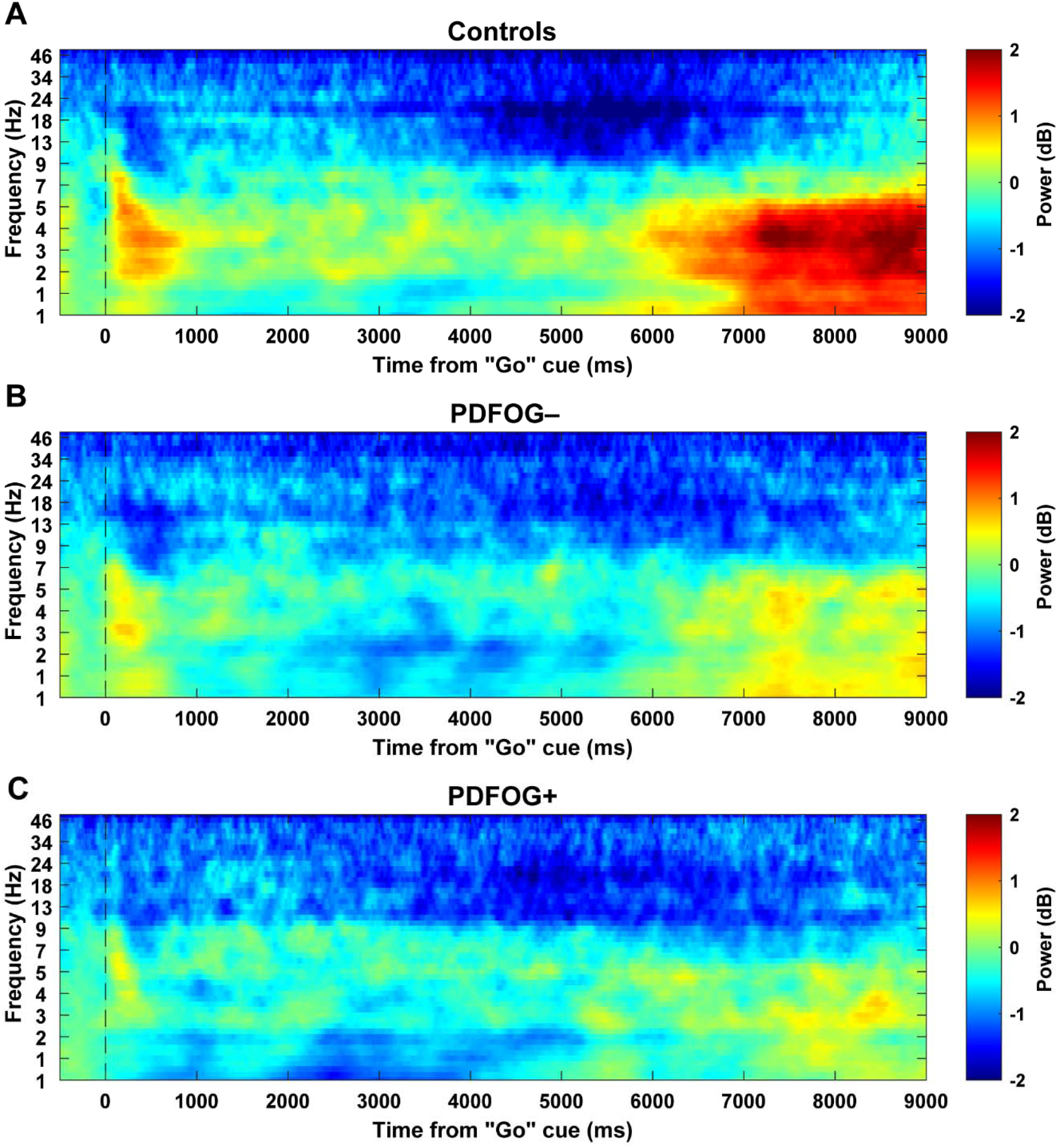
Time frequency distribution for the complete time window (−500 to 9000 ms around ‘Go’ cue) during the 7-second interval timing task. Time frequency plots for controls (A), PDFOG– (B), and PDFOG+ (C) during the interval timing task.

**Fig. S3.**
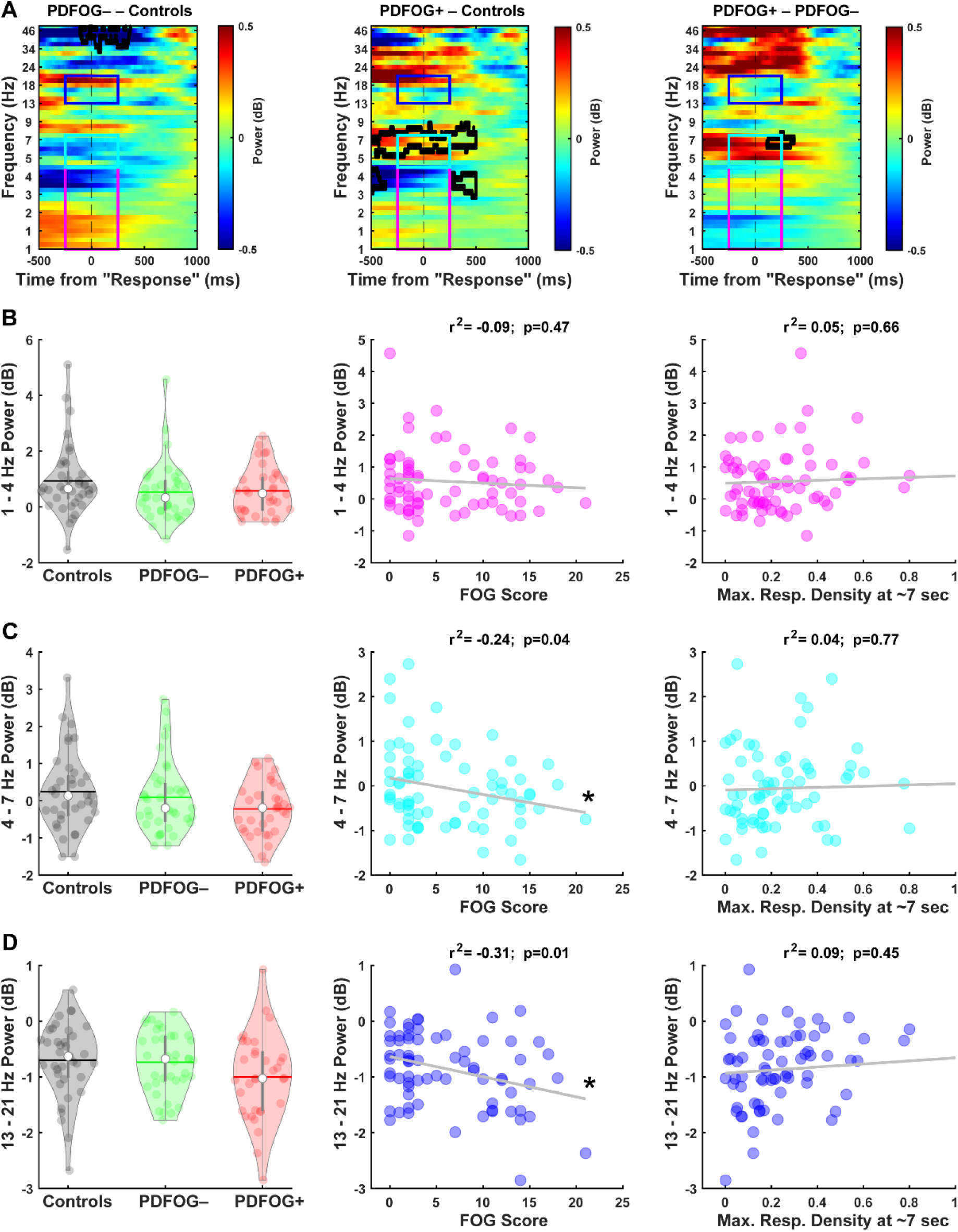
Response-triggered midfrontal delta, theta, and beta power values were not different in PDFOG+ during the interval timing task. (A) Time-frequency analyses comparing PDFOG+, PDFOG-, and controls around the key press (−500 to 1000 ms). (B) Response-triggered midfrontal delta power (1-4 Hz, tf-ROI: magenta box) was not different in the PDFOG+ group compared to other groups. Response-triggered midfrontal delta power in PD patients was not correlated with FOG scores as well as with maximum response density. (C) Response-triggered midfrontal theta power (4-7 Hz, tf-ROI: cyan box) was not different in the PDFOG+ group compared to other groups. Response-triggered mid-frontal theta power in PD patients was only correlated with FOG scores, but not with maximum response density. (D) Response-triggered midfrontal beta power (13-21 Hz, tf-ROI: blue box) was not different in the PDFOG+ group compared to other groups. Response-triggered mid-frontal beta power in PD patients was only correlated with FOG scores, but not with maximum response density. *p < 0.05 significant correlation. A-C: Areas outlined by solid black lines indicate p < 0.05 via a t-test.

